# Why estimating population-based case fatality rates during epidemics may be misleading

**DOI:** 10.1101/2020.03.26.20044693

**Authors:** Lucas Böttcher, Mingtao Xia, Tom Chou

## Abstract

Different ways of calculating mortality ratios during epidemics can yield widely different results, particularly during the COVID-19 pandemic. We formulate both a survival probability model and an associated infection duration-dependent SIR model to define individual- and population-based estimates of dynamic mortality ratios. The key parameters that affect the dynamics of the different mortality estimates are the incubation period and the length of time individuals were infected before confirmation of infection. We stress that none of these ratios are accurately represented by the often misinterpreted case fatality ratio (CFR), the number of deaths to date divided by the total number of infected cases to date. Using available data on the recent SARS-CoV-2 outbreaks and simple assumptions, we estimate and compare the different dynamic mortality ratios and highlight their differences. Informed by our modeling, we propose a more systematic method to determine mortality ratios during epidemic outbreaks and discuss sensitivity to confounding effects and errors in the data.

## 1. INTRODUCTION

The mortality ratio is a key metric describing the severity of a viral disease. It changes in time and can be measured in a number of ways during an epidemic. One common metric is the case fatality ratio (CFR), found by dividing the total number of deaths to date, *D*(*t*), by the total number of all cases to date *N*(*t*) [1, 2].

In the recent outbreaks of SARS-CoV-2, the CFR has been estimated from aggregated population data. We show examples of CFR curves in Fig. 1 and in the *Supplemental Information* (SI). As of March 26, 2020, the global CFR(*t*) = *D*(*t*)*/N*(*t*) = 21, 306*/*472, 762 ≈ 4.5% [3], while CFRs in individual regions vary significantly. Clearly, this estimate would correspond to an actual mortality ratio only if *all* the remaining unresolved individuals recover. However, some of these unresolved cases will lead to death and thus to a gradual increase of the estimated mortality ratio over time. Despite the underestimation of this type of population-based measurement, it is still commonly being used by various health officials and is often inconsistently defined as deaths/(deaths + recovereds) even though this difference has been clearly distinguished [4].

**FIG. 1.**
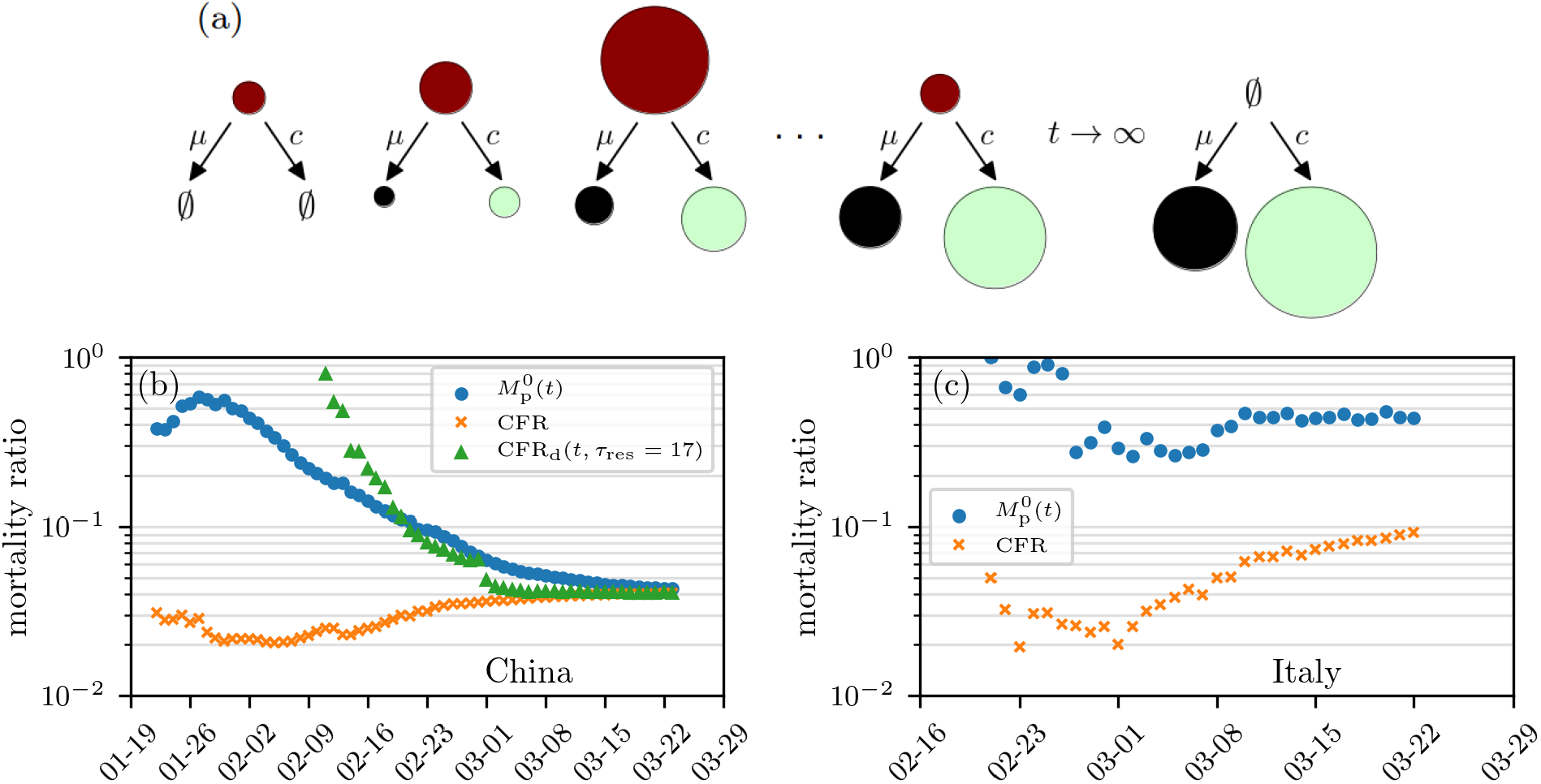
Mortality-ratio estimates. (a) Evolution of the cumulative number of infected (red), death (black), and recovered (green) cases. The size of the circles indicates the number of cases in the respective compartments on a certain day. (b–c) Estimates of mortality ratios (see Eqs. (8) and (14)) of SARS-CoV-2 infections in China and Italy. The “delayed” mortality-ratio estimate CFR_d_ corresponds to the number of deaths to date divided by total number of cases at time *t* − *τ*_res_. Many studies use CFR_d_, although this metric underestimates the individual-based mortality (defined below). Another population-based mortality ratio is *M*_p_(*t*), the number of deaths divided by the sum of death and recovered cases, up to time *t*. The data are based on Ref. [5].

During the severe 2003 acute respiratory syndrome (SARS) outbreak in Hong Kong, the World Health Organization (WHO) also used the aforementioned estimate to obtain a CFR of 4.5% while the final values approached 17.0% [6, 7]. For the ongoing SARS-CoV-2 outbreaks, analyses by WHO and other institutions still use the CFR = *D*(*t*)*/N*(*t*) metric (see Table I). Since actual mortality probabilities are important measures for assessing the risks associated with epidemic outbreaks, typical underestimations by CFRs may lead to insufficient countermeasures and a more severe epidemic [8, 9].

**TABLE I.** Different CFR estimates of COVID-19.

| reference | CFR |
| --- | --- |
| Xu *et al.* [1, 11] and Mahase [12] | 2 % |
| Wu *et al.* [2] | 0.1-1 % (outside Wuhan) |
| World Health Organization [13, 14] | 2-4 % |

An unambiguous, physiologically-based definition of mortality ratio is the probability that a single, newly infected individual will eventually die of the disease. If there are sufficient individual-level or cohort data, these probabilities can be further stratified according to patient age, gender, health condition, etc. [10]. The mortality ratio or probability of death should be an *intrinsic* property of the virus and the infected individual, depending on age, health, access to health care, etc. This intrinsic probability ought not to be *directly* dependent on the population-level dynamics of infected and recovered individuals. Thus, it can in principle be framed by a model for the survival probability of a single infected individual. Whether this individual infects others does not directly affect his probability of eventually dying. In section II A, we derive a model describing the probability *M*_1_(*t*) that an infected individual dies or recovers before time *t*. Importantly, these models incorporate the duration of infection (including an incubation period) before a patient tests positive at time *t* = 0.

However, as mentioned above, the CFR and other mortality measures are typically reported based on population data. Do these population-based measures, including CFR, provide reasonable measures of the probability of death of an individual? In section II B, we describe how mortality ratios are defined within population-level models, specifically, a disease duration-structured SIR model. We will show that population-based estimates are typically not a meaningful measure of mortality, but that under simplifying assumptions, the mortality ratio *M*_p_(*t*) is more closely related to the number of deaths to date divided by the number of dead plus the number of *recovered* individuals to date [4]. In the simplest approximation, the mortality ratio is currently (as of March 26, 2020) 21, 306*/*(21, 306+114, 749) ≈ 15.7% [3], significantly higher than the March 26, 2020 CFR ≈ 4.5% estimate.

We use the same estimates for the rate parameters in our individual and population models to compute the different mortality ratios. Note that in general, both the individual mortality probability *M*_1_(*t*) and the population-based estimates *M*_p_(*t*) depend on the time of measurement *t*. By critically analyzing these estimates and another *ad hoc* “delayed” ratio CFR_d_, we illustrate and interpret the differences among these measures and discuss how changes or uncertainty in the data affect them. In section III, we summarize our results and identify a correction factor to transform population-level mortality estimates into individual mortality probabilities.

## II. RESULTS

### A. Intrinsic individual mortality rate

Consider an individual that, at the time of positive testing (*t* = 0), had been infected for a duration *τ*_1_. A “survival” probability density can be defined such that *P*(*τ, t*|*τ*_1_)d*τ* is the probability that the patient is still alive and infected (not recovered) at time *t >* 0 and has been infected for a duration between *τ* = *t* + *τ*_1_ and *τ* + d*τ*. Since *τ*_1_ is unknown, it must be estimated or averaged over. The individual survival probability evolves according to

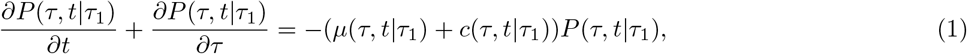

where the death and recovery rates, *µ*(*τ, t*| *τ*_1_) and *c*(*τ, t*| *τ*_1_), depend explicitly on the duration of infection at time *t* and implicitly on patient health and age *a* [15]. They may also depend explicitly on time *t* to reflect changes in clinical policy or available health care. For example, enhanced medical care may decrease the death rate *µ*, giving the individual’s intrinsic physiological processes a chance to cure the patient. These intrinsic individual-based death and recovery rates do not directly depend on population-level viral transmission.

Equation (1), assuming an initial condition of one particular individual who has been infected for time *τ*_1_ at the time of positive test, can be solved using the method of characteristics. From the solution *P*(*τ, t*| *τ*_1_) one can derive the probabilities of death and recovery by time *t* as

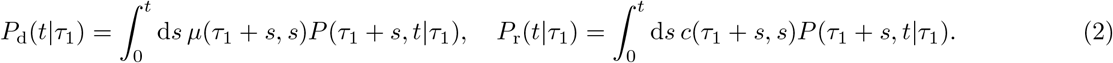

The probability that an individual died before time *t*, conditioned on resolution (either death or recovery), is then defined as

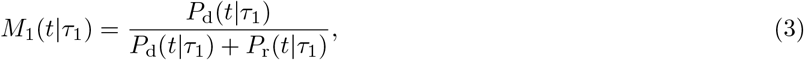

where we have explicitly indicated the dependence on the duration of the infection prior to confirmation of infection. These formulae also depend on all other relevant patient attributes such as age, accessibility to health care, etc. In the long-time limit, when resolution has occurred (*P*_d_(∞) + *P*_r_(∞) = 1), the individual mortality ratio is simply *M*_1_(∞) = *P*_d_(∞). This result relies only on intrinsic individual rate parameters and is completely independent of disease transmission at the population level. In order to capture the dependence of death and recovery rates on the time an individual has been infected, we propose a constant recovery rate *c* and a simple piece-wise constant death rate *µ*(*τ* |*τ*_1_) that is not explicitly a function of time *t*:

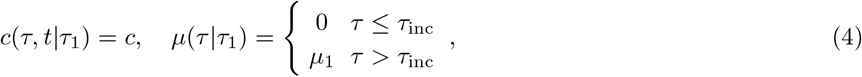

where *τ*_inc_ is the incubation-time parameter, the time after infection during which an individual remains asymptomatic. During this incubation period, the patient has zero death rate but can recover by clearing the virus. In other words, some patients fully recover without ever developing serious symptoms.

For coronavirus infections, the incubation period appears to be highly variable with a mean of *τ*_inc_ ≈ 6.4 days [17]. We can estimate *µ*_1_ and *c* using individual patient data where 19 patients (outside Hubei) had been tracked from the date on which their first symptoms occurred until the disease resolved [16].

Two out of 19 patients died, on average, 20.5 days after first symptoms occurred and the mean recovery time of the remaining 17 patients is 16.8 days. We show the recovery-time distribution in Fig. 2(a). Since we know that the mortality ratio in this dataset is 2*/*19, we can determine the dependence between *µ*_1_ and *c* according to *µ*_1_*/*(*µ*_1_ + *c*) ≈ 2*/*19 (or *c/µ*_1_ ≈ 8.5). The constant recovery and after-incubation period death rates [18] are estimated to be

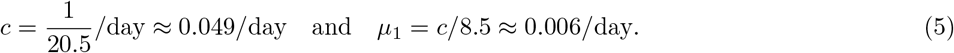

**FIG. 2.**
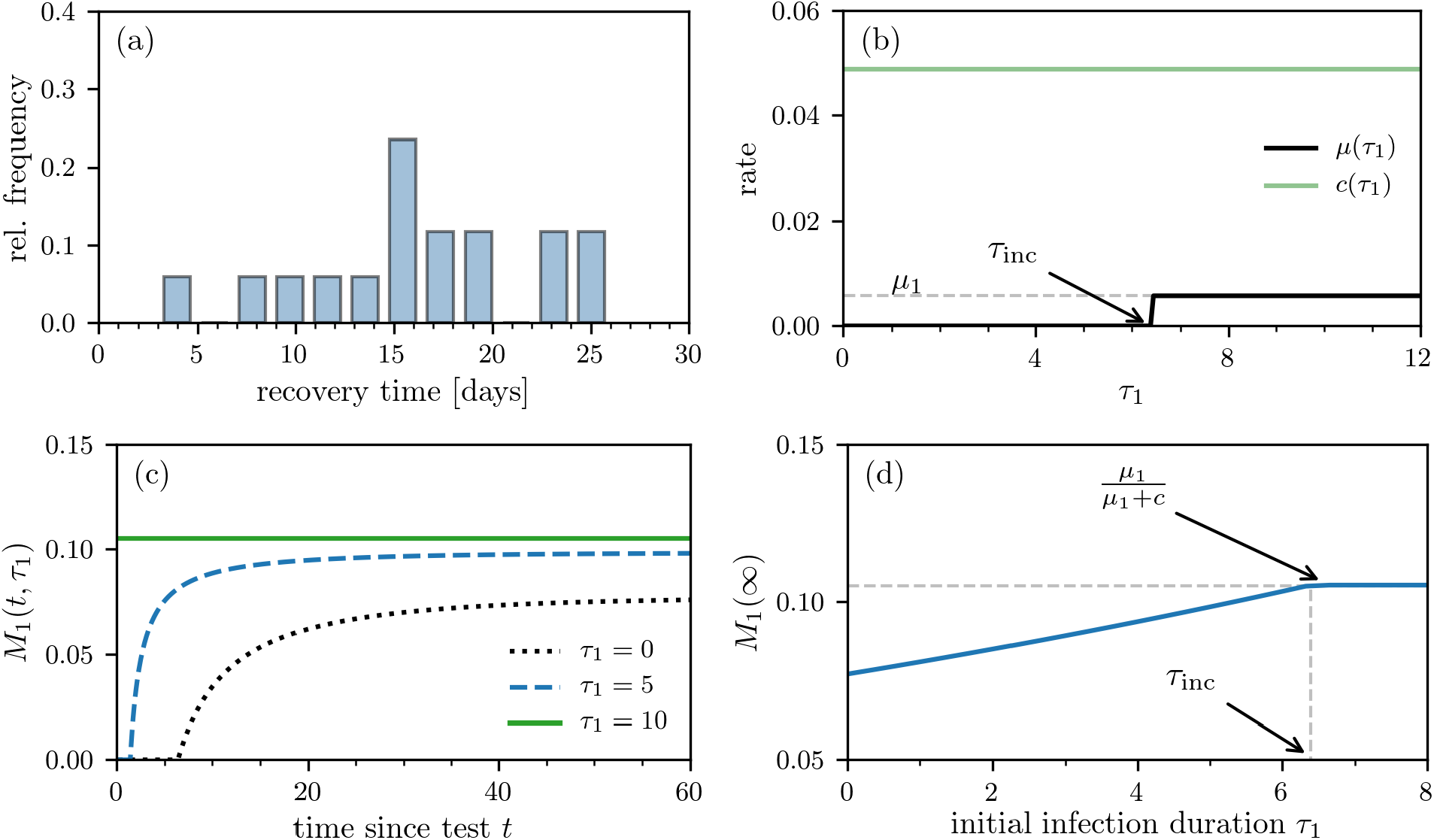
Individual mortality ratio. (a) Recovery time after first symptoms occurred based on individual data of 17 patients [16]. (b) Death- and recovery rates as defined in Eq. (4). The death rate *µ*(*τ*_1_) approaches *µ*_1_ for *τ*_1_ *> τ*_inc_, where *τ*_inc_ is the incubation period and *τ*_1_ is the time the patient has been infected before first being tested positive. (c) The individual mortality ratio *M*_1_(*t* |*τ*_1_) for *τ*_inc_ = 6.4 days at different values of *τ*_1_. Note that the individual death probability *P*_d_(*t* |*τ*_1_) and *M*_1_(*t*| *τ*_1_) are nonzero only after *t > τ*_inc_− *τ*_1_. (d) The asymptotic individual mortality ratio *M*_1_(∞) (see Eq. (3)) as a function of *τ*_1_.

Using these numbers, the recovery and death rate functions *c*(*τ, t*| *τ*_1_) and *µ*(*τ* |*τ*_1_) are plotted as functions of *τ* in Fig. 2(b). We show the evolution of *M*_1_(*t* |*τ*_1_) at different values of *τ*_1_ in Fig. 2(c). The corresponding long-time limit *M*_1_() is readily apparent in Fig. 2(d): for *τ*_1_ ≥ *τ*_inc_, *M*_1_(∞) = *µ*_1_*/*(*µ*_1_ +*c*) ≈ 0.105, while *M*_1_(∞) *< µ*_1_*/*(*µ*_1_ +*c*) when *τ*_1_ *< τ*_inc_. The smaller expected mortality associated with early identification of infection arises from the remaining incubation time during which the patient has a chance to recover without possibility of death. When conditioned on testing positive at or after the incubation period, the patient immediately suffers a positive death rate, increasing his *M*_1_(∞).

Finally, in order to infer *M*_1_ (and also indirectly *µ* and *c*) during an outbreak, a number of statistical issues must be considered. First, if the outbreak is ongoing, there may not be sufficient long-time cohort data. Second, *τ*_1_ is unknown. Since testing typically occurs at the onset of symptoms, most positive patients will have been infected a few days earlier. The uncertainty in *τ*_1_ can be represented by a probability density *ρ*(*τ*_1_) for the individual. The expected mortality can then be constructed as an average over *ρ*(*τ*_1_):

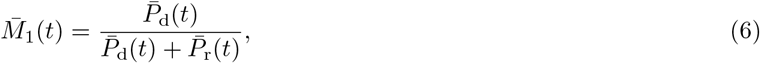

where 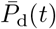 and 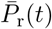 are the the *τ*_1_-averaged probabilities death and cure probabilities. Note that this averaging is different from the population-level averaging 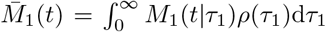, which would describe the average of mortality ratios over a population with heterogeneous initial durations *τ*_1_.

Some properties of the distribution *ρ*(*τ*_1_) can be inferred from the behavior of patients. Before symptoms arise, only very few patients will know they have been infected, seek medical care, and get their case confirmed (*i*.*e*., *ρ*(*τ*_1_) ≈ 0 for *τ*_1_ ≈ 0). The majority of patients will contact hospitals/doctors when they have been infected for a duration of *τ*_inc_. The distribution *ρ*(*τ*_1_) thus reaches its maximum near or shortly after *τ*_inc_. Since patients are most likely to test positive after experiencing symptoms, we choose a gamma distribution

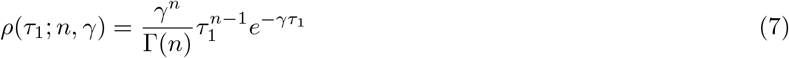

with shape parameter *n* = 8 and rate parameter *γ* = 1.25*/*day so that the mean *n/γ* is equal to *τ*_inc_ = 6.4.

Upon using the rates in Eqs. (4) and averaging over *ρ*(*τ*_1_), we derived expressions for 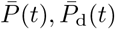, and 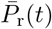 which are explicitly given in the SI. Using the values in Eq. (5) we find an expected individual mortality ratio 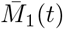 (which are subsequently plotted in Fig. 3) and its asymptotic value 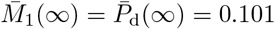. Of course, it is also possible to account for more complex time-dependent forms of *c* and *µ*_1_ [19], but we will primarily use Eqs. (4) in our subsequent analyses.

**FIG. 3.**
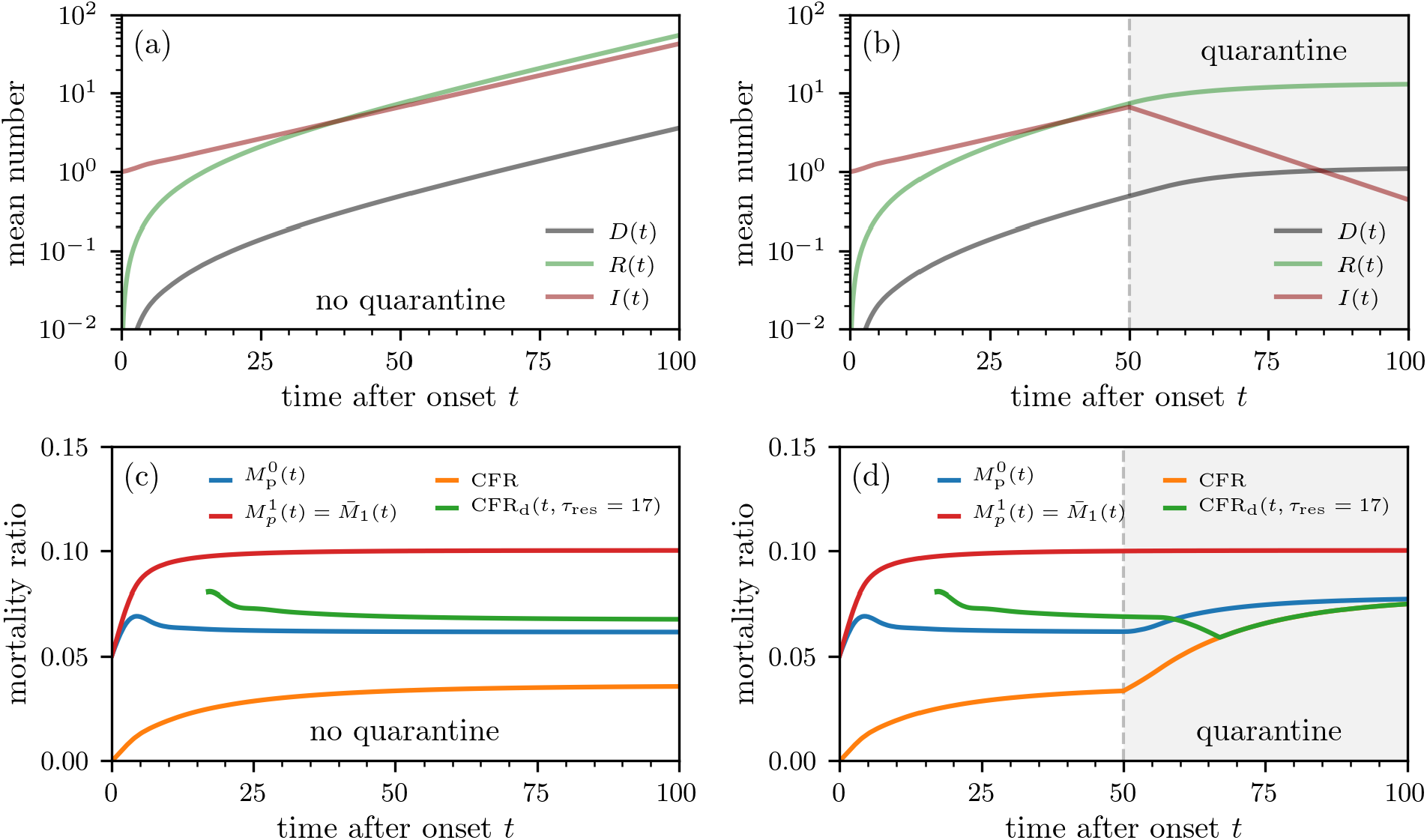
Population-level mortality-ratio estimates. Outbreak evolution and mortality ratios without containment measures (a,c) and with quarantine (b,d). The curves are based on numerical solutions of Eqs. (9) using the initial condition *I*(*τ*, 0) = *ρ*(*τ* ; 8, 1.25) (see Eq. (7)). The death and recovery rates are defined in Eqs. (4) and (5). We use a constant infection rate *β*_1_*S*(0) = 0.158/day, which we estimated from the basic reproduction number of SARS-CoV-2 [17]. To model quarantine effects, we set *β*_1_ = 0 for *t >* 50. We show the mortality-ratio estimates 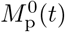 and 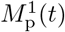 (see Eq. (14)) and CFR_d_(*t, τ*_res_) (see Eqs. (8), (11), (12), and (14)).

In the next section, we define population-based estimates for mortality ratios, *M*_p_(*t*), and explore how they can be computed using SIR-type models. By comparing *M*_1_(*t*) to *M*_p_(*t*), we gain insight into whether population-based metrics are good proxies for individual mortality ratios. We will outline the mathematical differences and additional errors that confound population-level estimates.

### B. Infection duration-dependent SIR model

While individual mortalities can be estimated by tracking many individuals from infection to recovery or death, oftentimes, the available data are not resolved at the individual level and only total populations are given. Typically, one has the total number of cases accumulated up to time *t, N*(*t*), the number of deaths to date *D*(*t*), and the number of cured/recovered patients to date *R*(*t*) (see Fig. 1). The CFR is simply *D*(*t*)*/N*(*t*). Note that *N*(*t*) includes unresolved cases and that *N*(*t*) ≥ *R*(*t*)+ *D*(*t*). Resolution (death or recovery) of all patients, *N* (∞) = *R*(∞)+ *D*(∞), occurs only well after the epidemic passes.

A variant of the CFR commonly used in the literature [1, 2] is the delayed CFR

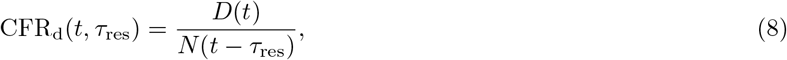

where *τ*_res_ is a corresponding time lag that accounts for the duration from the day when first symptoms occurred to the day of cure/death. Many estimates of the COVID-19 mortality ratio assume that *τ*_res_ = 0 [1, 2] and thus underestimate the number of death cases *D*(*t*) that result from a certain number of infected individuals. Similar underestimations using CFR_d_ have been reported in previous epidemic outbreaks of SARS [4, 6] and Ebola [20].

Alternatively, a simple and interpretable population-level mortality ratio is *M*_p_(*t*) = *D*(*t*)*/*(*R*(*t*) + *D*(*t*)), the death ratio of all *resolved* cases. To provide a concrete model for *D*(*t*) and *R*(*t*), and hence *M*_p_(*t*), we will use a variant of the standard infection duration-dependent susceptible-infected-recovered (SIR)-type model described by [21]

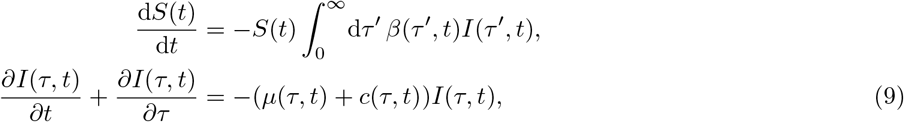

and 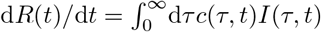, where *S*(*t*) is the number of susceptibles, *I*(*τ, t*) is density of individuals at time *t* who have been infected for time *τ*, and *R*(*t*) is the number of recovered individuals. The rate at which an individual infected for time *τ* at time *t* transmits the infection to a susceptible is denoted by *β*(*τ, t*)*S*(*t*).

Note that the equation for *I*(*τ, t*) is identical to the equation for the survival probability described by Eq. (1). It is also equivalent to McKendrick age-structured models [22, 23] [24]. Infection of susceptibles is described by the boundary condition

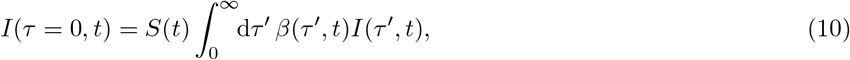

which is similar to that used in age-structured models to represent birth [22]. Finally, we use an initial condition consistent with the infection duration density given by Eq. (7): *I*(*τ*, 0) = *ρ*(*τ* ; *n* = 8, *γ* = 1.25). Note that Eq. (10) assumes that all newly infected individuals are immediately identified; *i*.*e*., these newly infected individuals start with *τ*_1_ = 0. After solving for the infected population density, the total number of deaths and recoveries to date can be found via

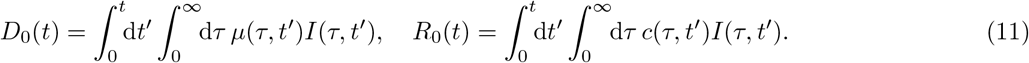

The corresponding total number of cases *N*(*t*) in Eq. (8) is

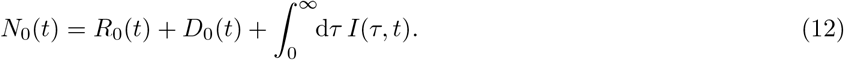

In the definitions of *D*_0_(*t*), *R*_0_(*t*), and *N*_0_(*t*), we account for all possible death and recovery cases to date (see SI) and that newly infected individuals are immediately identified. We use these case numbers as approximations of the reported case numbers to study the evolution of mortality-ratio estimates. Mortality ratios based on these numbers underestimate the actual individual mortality *M*_1_ (see section II A) since they involve individuals that have been infected for different durations *τ*, particularly recently infected individuals who have not yet died.

An alternative way to compute populations is to exclude the newly infecteds and consider only the initial cohort. The corresponding populations in this case are defined as

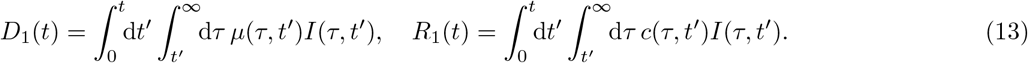

Since *D*_1_(*t*) and *R*_1_(*t*) do not include infecteds with *τ < t*, they exclude the effect of newly infected individuals, but may yield more accurate mortality-ratios as they are based on an initial cohort of individuals in the distant past. The infections that occur after *t* = 0 contribute only to *I*(*τ < t, t*); thus, *D*_1_(*t*) and *R*_1_(*t*) do not depend on the transmission rate *β* or the number of susceptibles *S*(*t*). Note that all the populations derived above implicitly average over *ρ*(*τ*_1_; *n, γ*) for the first cohort of identified infecteds (but not subsequent infecteds). Moreover, the population density *I*(*τ ≥ t, t*) follows the same equation as 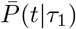 provided the same *ρ*(*τ*_1_; *n, γ*) is used in their respective calculations.

The two different ways of partitioning populations (Eqs. (11) and (13)) lead to two different population-level mortality ratios

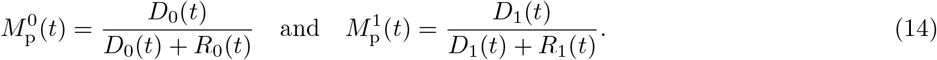

Since the populations *D*_0_(*t*) and *R*_0_(*t*), and hence 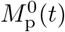, depend on disease transmission through *β* (*τ*; *t*) and *S*(*t*), we expect 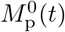 to carry a different interpretation from M_1_(*t*) and 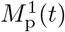.

In the special case in which *µ* and *c* are constants, the time-integrated populations 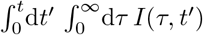 and 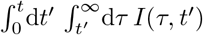 factor out of 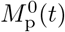 and 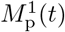, rendering them time-independent and

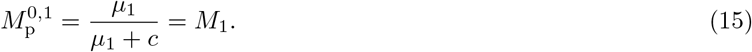

Thus, only in the special time-homogeneous case do both population-based mortality ratios become *independent* of the population (and transmission *β*) and coincide with the individual death probability.

To illustrate the differences between 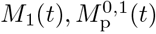, and CFR_d_(*t, τ*_res_) in more general cases, we use the simple death and cure rate functions given by Eqs. (4) in solving Eqs. (1) and (9). For *β*(*τ, t*) in Eq. (10), we account for incubation effects by neglecting transmission during the asymptomatic incubation period (*τ* ≤ *τ*_inc_) and assume

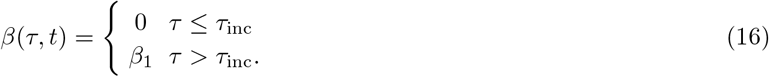

We use the estimated basic reproductive number ℛ_0_ = *β*_1_*S*(0)*/*(*µ*_1_ + *c*) ≈ 2.91 [17] to fix *β*_1_*S*(0) = (*µ*_1_ + *c*) ℛ_0_ ≈ 0.158*/*day. We also first assume that the susceptible population does not change appreciably before quarantine and set *S*(*t*) = *S*(0). Thus, we only need to solve for *I*(*τ, t*) in Eqs. (9) and (10). We solve Eqs. (9) and (10) numerically (see the *Methods* section for further details) and use these numerical solutions to compute *D*_0,1_(*t*), *R*_0,1_(*t*), and *N*_0,1_(*t*) (see Fig. 3(a) and (b)), which are then used in Eqs. (14) and CFR_d_(*t* − *τ*_res_). To determine a realistic value of the time lag *τ*_res_, we use data on death/recovery periods of 36 tracked patients [16] and find that patients recover/die, on average, *τ*_res_ = 16.5 days after first symptoms occurred.

We show in Figs. 3(c) and (d) that 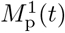 approaches the individual mortality ratio 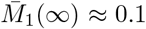 of section II A. This occurs because the model for *P*(*τ, t*) and *I*(*τ, t*) are equivalent and we assumed that the initial distribution of *τ* for both quantities are given by *ρ*(*τ* ; 8, 1.25). However, the population-level mortality ratios CFR_d_(*t, τ*_res_) and 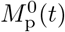 also take into account recently infected individuals who may recover before symptoms. This difference yields different mortality ratios because newly infecteds are implicitly assumed to be detected immediately and all have *τ*_1_ = 0. Thus, the underlying infection-time distribution is not the same as that used to compute 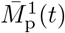 (see SI for further details). The mortality ratios CFR_d_(*t, τ*_res_) and 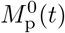 should not be used to quantify the individual mortality probability of individuals who tested positive after their incubation period. During the course of an outbreak, the measures CFR_d_(*t, τ*_res_) and 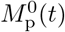 are subject to another confounding influence. Since *D*(*t*), *R*(*t*), and *N*(*t*) do not change with the same rates at the same time, these population-level mortality estimates only reach their steady state after sufficiently long times (see Fig. 3(c) and (d)).

To summarize, we described two confounding factors that complicate the direct use of population-level mortality ratio to estimate individual mortality probabilities. First, infection-time distributions *ρ*(*τ* ; *n, γ*) that are meaningful on an individual level may not correspond to those in population-level data. Second, population-level mortality ratios are often time-dependent and most informative only in the steady state after the outbreak stopped.

The evolution of the mortality ratios in Fig. 3 qualitatively resembles the behavior of the mortality-ratio estimates in Fig. 1. As shown in Fig. 1, the population-based estimates for coronavirus varies, decreasing in time for China but fluctuating for Italy. These changes could result from changing practices in data collecting, or from explicitly time-inhomogeneous parameters *µ*(*τ, t*), *c*(*τ, t*), and/or *β*(*τ, t*).

Although population-level quarantining does not directly affect the individual mortality *M*_1_(*t* | *τ*_1_) or 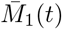, it can be easily incorporated into the SIR-type population dynamics equations through changes in *β*(*τ, t*)*S*(*t*). For example, we have set *S*(*t > t*_q_) = 0 to represent implementation of a quarantine after *t*_q_ = 50 days of the outbreak. After *t*_q_ = 50 days, no new infections occur and the estimates CFR_d_(*t, τ*_res_) and 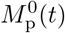 start converging immediately towards their steady-state values (see Fig. 3(d)). Since the number of deaths decreases after the implementation of quarantine measures, the delayed CFR_d_(*t, τ*_res_ = 17) is first decreasing until *t* = *t*_q_ + *τ*_res_ = 67. For *t >* 67, the CFR_d_(*t, τ*_res_ = 17) measures no new cases and is thus equal to the CFR.

## III. DISCUSSION AND SUMMARY

After an outbreak, it is important to assess the severity of the disease by estimating its mortality and other disease characteristics. Assuming accurate data, the often-used CFR and delayed CFR typically underestimate the true, final death ratio. For example, during the SARS outbreaks in Hong Kong, the WHO first estimated the fatality rate to 2.5% (March 30, 2003) whereas the final estimates reached values of about 17.0% (June 30, 2003) [7]. Standard metrics like the CFR are seen to be easily confounded by and sensitive to uncertainty in intrinsic disease parameters such as the incubation period and the time *τ*_1_ a patient had been infected before clinical confirmation of infection. For the recent COVID-19 outbreaks, CFR-based measures may still provide reasonable estimates of the actual mortality across different age classes due to a counter-acting error in the numbers of unreported mild-symptom cases.

Here, we stress that more mechanistically meaningful and interpretable metrics can be defined and be as easily estimated from data as CFRs. Our proposed mortality ratios for viral epidemics are defined in terms of (i) individual survival probabilities and (ii) population ratios using numbers of deaths and recovered individuals. Both of these measures are based on the within-host evolution of the disease, and in the case of 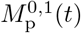, the population-level transmission dynamics. Thus, these metrics directly incorporate key parameters operating on the weeks or months timescale, the incubation time *τ*_inc_ and time of prior infection *τ*, through the solution of age-structured PDEs. Among the metrics we describe, 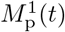 is structurally closest to 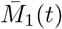 in that both are independent of transmission *β* since new infections are not considered. Both of these converge after an incubation time *τ*_inc_ to a value smaller than or equal to *µ*_1_*/*(*µ*_1_ + *c*).

The most accurate estimates of *M*_1_ can be obtained if we keep track of the fate of cohorts that were infected within a small time window in the past. By following only these individuals, one can track how many of them died as a function of time. As more cases arise, one should stratify them according to estimated *τ* to gather improving statistics for *M*_1_(∞). These data should also be collated according to the other central factor in COVID-19 mortality: patient age. With the further spread of SARS-CoV-2 in different countries, data on more individual cases of death and recovery can be more easily stratified by age, health condition, and other individual characteristics. Using identical initial infection time distributions *ρ*(*τ*_1_; *n, γ*) (see Eq. (7)), the long-time limit of 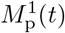 approaches the individual mortality 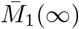 (see Eq. (6)).

Besides accurate cohort data, for which at present there are few for coronavirus, cumulative population data has been used to estimate the mortality ratio. The metrics 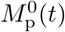 and CFR(*t*) are based on these aggregate populations but implicitly depend on new infections and the transmission rate *β*. Despite this confounding factor, 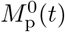 and CFR_d_(*t, τ*_res_) approach 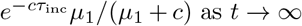 as *t* → ∞, where 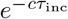 is the probability that no recovery occurred during the incubation time *τ*_inc_. Based on these results, we can establish the following connection between the different mortality ratios for initial infection times with distribution *ρ*(*τ*_1_; *n, γ*) and mean *τ* = *n/γ*:

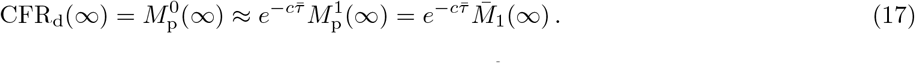

According to Eq. (17), population-level mortality estimates (*e*.*g*., CFR and 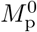) can be transformed, at least approximately, into individual mortality probabilities using the correction factor 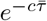 with 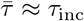.

Besides the mathematical differences between *M*_1_(*t*) and 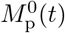, CFR, estimating 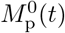 and CFR(*t*) from aggregate populations implicitly incorporate a number of confounding factors that lead to variability in these estimates. In Fig. 4, we plot the population-level mortality-ratio estimates 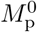 against the CFR for different regions and observe large variations and very little correlation between countries [25]. As of March 26, 2020, the value of 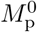 in Italy is almost 45% and can increase further if the current conditions (e.g., treatment methods, age group proportion of infecteds, etc.) do not change. Differences between the mortality ratios in China and Italy (see Figs. 1(b) and (c)) might be a result of varying medical treatment strategies, different practices in data collecting (e.g., post-mortem testing), and differences in the age demographics between the countries.

**FIG. 4.**
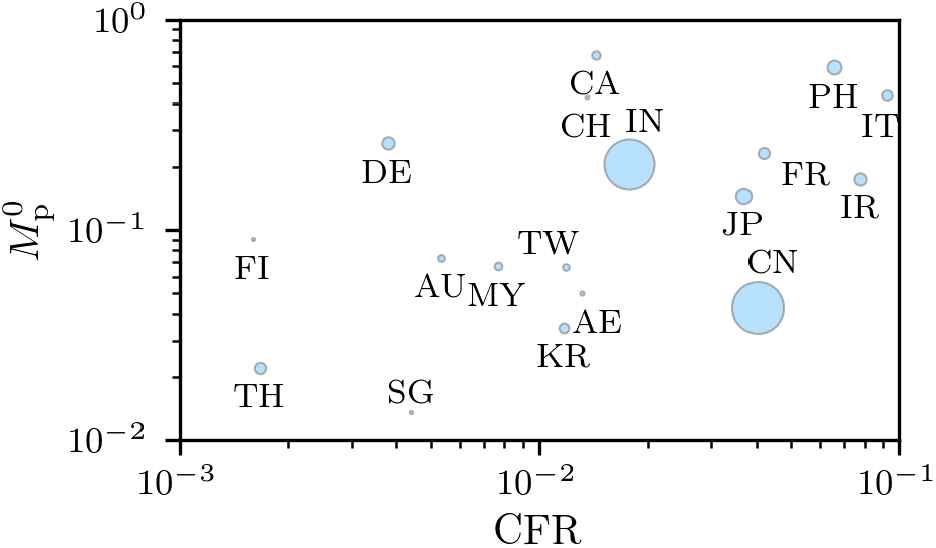
Region-dependence of COVID-19 mortality-ratio estimates. Mortality-ratio estimates of COVID-19 in different regions (see Eqs. (8) and 14 (*τ*_res_ = 0)). We used data on the cumulative number of cases, recoveries, and deaths in Ref. [5] as of March 24, 2020. The marker sizes indicate the population of the corresponding countries. The metrics 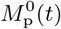 and and CFR are largely uncorrelated with correlation coefficient 0.33.

In general, even if the cohort initially tested was only a fraction of the total infected population, tracking 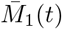 or 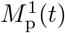 of this cohort still provides an accurate estimation of the mortality rate. However, the newly infecteds that contribute to CFR and 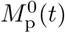 at later times may not all be tested or may be tested at different times after they were infected. A reported/tested fraction *f <* 1 *would not* directly affect the CFRs or mortality ratios if the unreported/untested individuals die and recover in the same proportion as the tested infecteds. Undertesting will overestimate true CFR or mortality rates if the untested infecteds are less likely to die than the tested infecteds. In other words, if the untested (presumably because they were mildly or asymptomatic) population predominantly recovers instead of dying, the actual CFR and mortality ratios would be significantly lower than those based on tested individuals. If untested infecteds do not die, the asymptotic mortality of all infected individuals 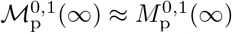 (see the SI). Current estimates show that only a minority of SARS-CoV-2 infections are reported (e.g., *f* ≈ 14% in China before January 23, 2020) [26].

Besides under-reporting, the delay in transmission after becoming infected will also affect 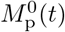. Although we have assumed that transmission occurs only after the incubation period when symptoms arise, there is evidence of asymptomatic transmission of coronavirus [26, 27]. Asymptomatic transmission can be modeled by setting *β*(*τ*) *>* 0 even for *τ < τ*_inc_. An undelayed transmission in a nonquarantine scenario causes relatively more new infecteds who have not had the chance to die yet, leading to a *smaller* mortality ratio 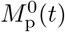. Within our SIR model, delaying transmission reduces the number of infected individuals and deaths at any given time but *increases* the measured mortality ratio 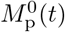. Without quarantine, the asymptotic values 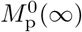 and CFR(∞) will also change as a result of changing the transmission latency period, as shown in the SI. With perfect quarantining, the asymptote 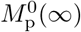 is eventually determined by a cohort that does not include new infections and is thus independent of the transmission delay.

In this work, we have explicitly defined a number of interpretable mathematical metrics that represent the risk of death. By rigorously defining these metrics, we are able to reveal the inherent assumptions and factors that affect their estimation. Within survival probability and SIR-type models, we explicitly illustrate how physiologically important parameters such as incubation time, death rate, cure rate, and transmissibility influence the metrics. We also discussed how statistical factors such as time of testing after infection (*τ*_1_) and testing ratio (*f*) affect our estimates. Given the uncertainty in the testing fraction, we conclude that *M*_1_(*t*) and 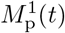 is best interpreted as approximately the mortality probability conditioned on being tested positive. In practice, these are probably also good estimates of mortality of patients conditioned on showing symptoms. In addition to our metrics and mathematical models, we emphasize the importance of curating individual cohort data. These data are more directly related to the probability of death *M*_1_(*t*) and are subject to the fewest confounding factors and statistical uncertainty.

## METHODS

To numerically solve Eqs. (9) and (10), we used a uniform discretization *τ*_*k*_ = *k*Δ*τ, k* = 0, 1, …, *K*. A backward difference operator [*I*(*τ*_*k*_, *t*) − *I*(*τ*_*k*−1_, *t*)] */*(Δ*τ*) is used to approximate ∂_*τ*_ *I*(*τ, t*) and a predictor-corrector Euler scheme is used to advance time [28]. Setting the cut-off *I*(−Δ*τ, t*) ≡ 0 and *I*(*K*Δ*τ, t*) ≡ 0, the resulting discretized equations for the full SIR model are

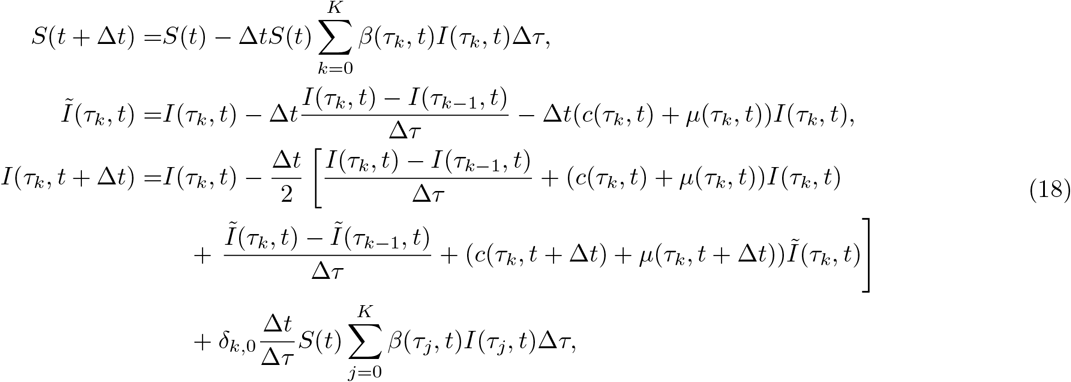

where 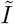 is the initial predicted guess, and the last term proportional to *δ*_*k*,0_ encodes the boundary condition Eq. (10). Note that we use 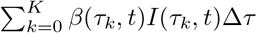 to indicate the numerical evaluation of 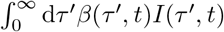. Quadrature methods such as Simpson’s rule and the trapezoidal rule can be used to approximate the integral more efficiently.

The total deaths, recovereds, and infecteds at time *t* are found by

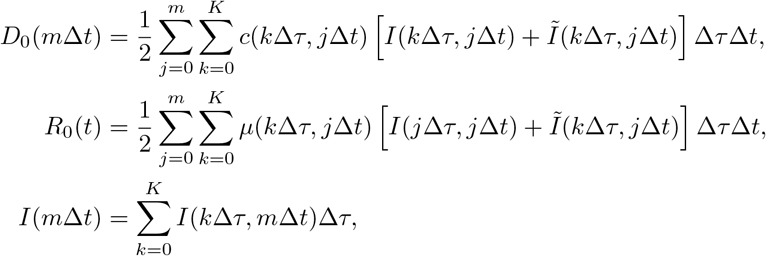

with analogous expressions for *D*_1_(*m*Δ*t*) and *R*_1_(*m*Δ*t*). To obtain a stable integration scheme, the time steps Δ*t* and Δ*τ* have to satisfy Δ*t*/(2Δ*τ*) < 1. In all of our numerical computations, we thus set Δ*t* = 0.002, Δ*τ* = 0.02, and *K* = 10^4^. In the SI, we show additional plots of the magnitude of *I*(*τ, t*) in the *t* − *τ* plane.

## Data Availability

The datasets that we used in this study are stored in the publicly accessible repositories of Refs. [3, 5, 16].

## ACKNOWLEDGEMENTS

LB acknowledges financial support from the SNF Early Postdoc.Mobility fellowship on “Multispecies interacting stochastic systems in biology”. The authors also acknowledge financial support from the Army Research Office (W911NF-18-1-0345), the NIH (R01HL146552), and the National Science Foundation (DMS-1814364).

## AUTHOR CONTRIBUTIONS

LB and TC developed the analyses and wrote the manuscript. LB analyzed data and MX performed the numerical computations.

## SUPPLEMENTARY INFORMATION

### A. Additional examples of mortality-ratio evolutions

**FIG. S1.**
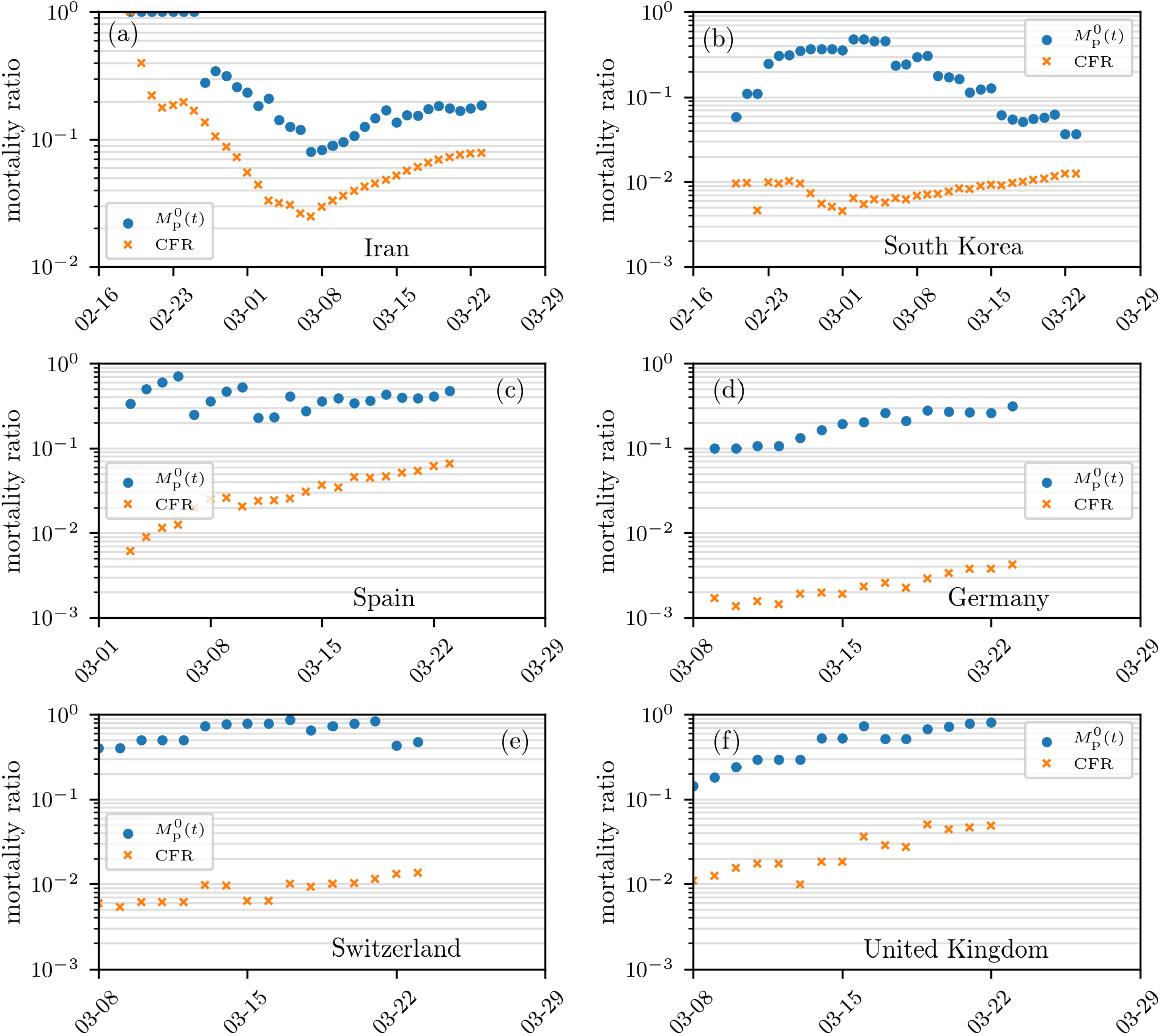
Mortality ratio estimates. Estimates of mortality ratios (see Eqs. (8) and (14) in the main text) of SARS-CoV-2 infections in different countries. The case fatality ratio, CFR, corresponds to the number of deaths to date divided by the total number of cases to date. Another population-based mortality ratio is 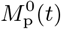, the number of deaths divided by the sum of deaths and recovereds, up to time *t*. The data are derived from Ref. [5].

In Fig. S1, we show additional examples of mortality-ratio estimates for Iran, South Korea, Spain, Germany, Switzerland, and the United Kingdom. As in Fig. 1 in the main text, we observe that, by definition, the population-based mortality ratio 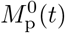 is significantly larger than the corresponding CFR in all cases.

### B. Solutions for *τ*_1_-averaged probabilities

Using the method of characteristics, we find the formal solution to Eq. (1):

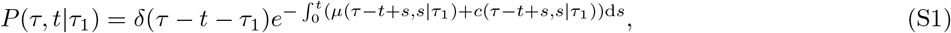

which can be used to construct the death and cure probabilities

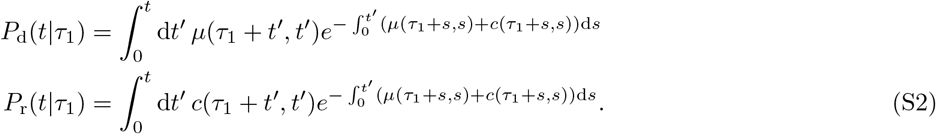

If we now invoke the functional forms of *µ* and *c* given in Eq. (4), we find explicitly

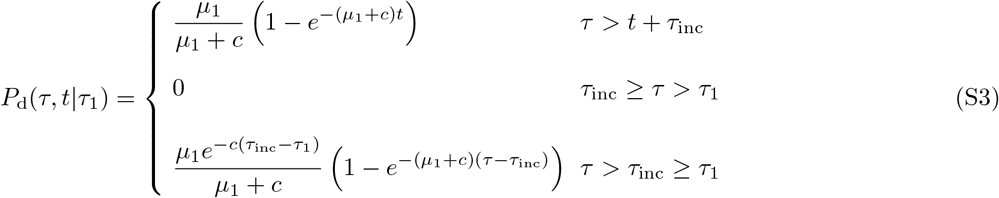

and

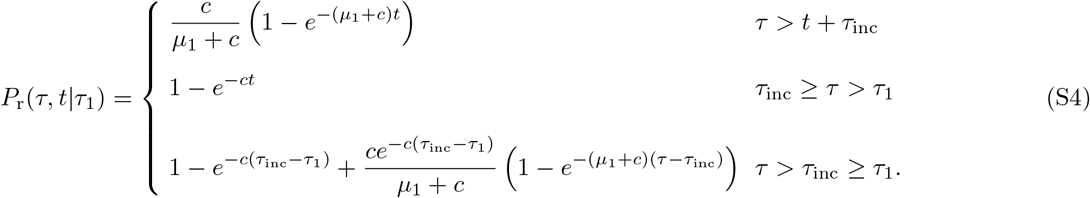

**FIG. S2.**
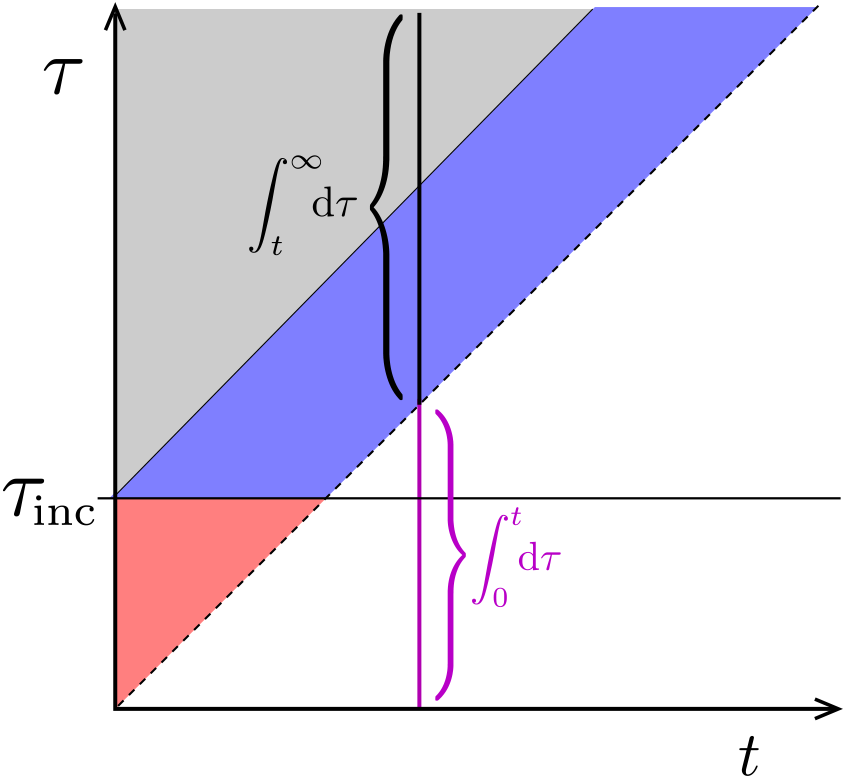
Phase plot for *P*(*τ > t, t*) and *I*(*τ > t, t*). The regions delineating different forms for the solution (Eq. (S5)). Here, we have included an incubation time *τ*_inc_ before which no death occurs. The solution for 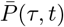 or *I*(*τ, t*) in the *τ < t* region must be self-consistently solved using the boundary condition Eq. (10). At any fixed time, the integral of *I*(*τ, t*) over *t < τ* ≤ ∞ captures only the initial population, excludes newly infecteds, and is used to compute *D*_1_(*t*), *R*_1_(*t*), and 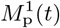. To compute *D*_0_(*t*), *R*_0_(*t*), and 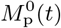, we integrate across all infecteds (including the integral over *t > τ >* ≥ 0 shown in magenta).

Finally, we can also find the *τ*_1_-averaged probabilities for *τ* ≥ *t* by weighting over *ρ*(*τ*_1_; *n, γ*). For example,

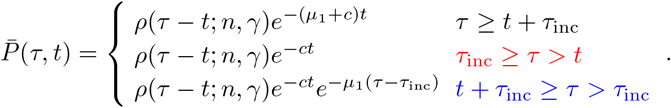

**FIG. S3.**
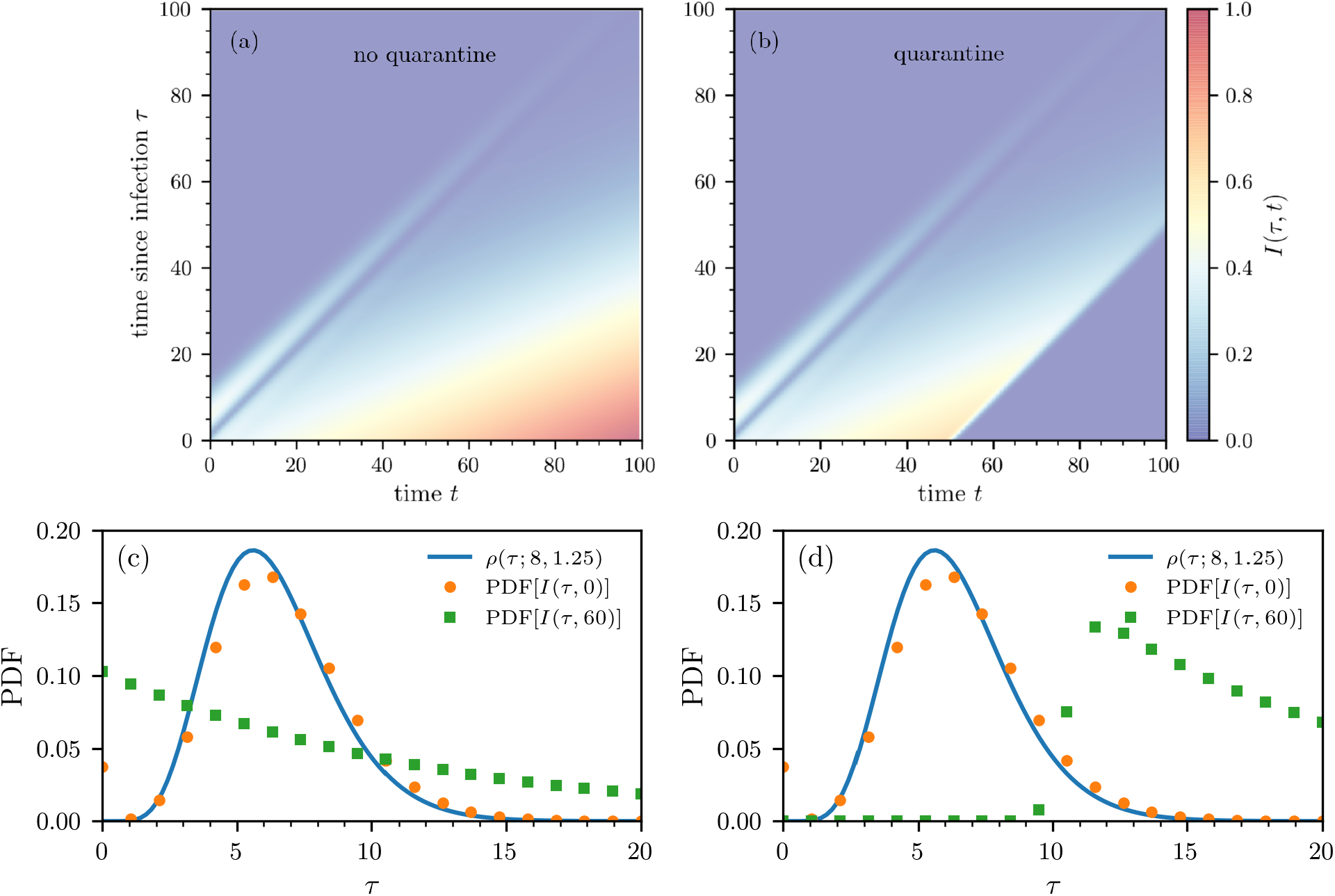
Density plots of *I*(*τ, t*) in the *t*− *τ* plane. Numerical solution of the equation for *I*(*τ, t*) in Eqs. (9) under the assumption of a fixed susceptible size *β*_1_*S* = 0.158/day. (a) The density without quarantine monotonically grows with time *t* in the region *τ < t* as an unlimited number of susceptibles continually produces infecteds. (b) With quarantining after *t*_q_ = 50 days, we set *β*_1_*S* = 0 for *t > t*_q_, which shuts off new infections. Both plots were generated using the same initial density *ρ*(*τ*_1_) defined in Eq. (7). In both cases, the density *I*(*τ > t*) is identical to *P*(*τ > t*) if the same *ρ*(*τ*_1_) is used and is independent of disease transmission, susceptible dynamics, etc. (c-d) Probability-density functions (PDFs) of the number of infected *I*(*τ, t*) for *t* = 0, 60 (b) without and (c) with quarantine. The blue solid line corresponds to the initial distribution *ρ*(*τ* ; *n* = 8, *γ* = 1.25) (see Eq. (7)).

These solutions hold for the different regions shown in the phase plot of Fig. S2 and are equivalent to those for *I*(*τ > t, t*). Corresponding expressions for 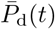 and 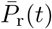 can be found and used to construct 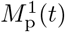. Fig. S3(a) shows the magnitude of *I*(*τ, t*) in the *t* − *τ* plane when we set *S*(*t*) = *S* constant (so that the first equation in Eq. (18) does not apply) such that *β*_1_*SS* ≈ 0.158/day. In this case, the epidemic continues to grow in time, but the mortality rates 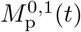 nonetheless converge as *t*. In Fig. S3(b), we set *β*_1_*S* = 0 for *t > t*_q_ to model strict quarantining after *t*_q_ = 50 days. We observe no new infections after the onset of strict quarantine measures. In both cases (quarantine and no quarantine), we use *ρ*(*τ* ; *n* = 8, *γ* = 1.25) (see Eq. (7) in the main text) to describe the initial distribution of infection times *τ*. As time progresses, more of the distribution of *τ* moves towards smaller values until quarantine measures take effect (see Fig. S3(c) and (d)).

### C. Effects of undertesting

Note that *I*(*τ, t*) in the SIR equations determines the dynamics of the actual infected population. However, (i) typically only a fraction *f* of the total number of infecteds might be tested and confirmed positive and (ii) the testing of newly infecteds may also be delayed by a distribution *ρ*(*τ* ; *n, γ*).

If positive tests represent only a fraction *f* of the total infected population, and the confirmation of newly infecteds occurs immediately, the known infected density is given by *I*^*∗*^(*τ, t*) = *f I*(*τ, t*) where *I*(*τ, t*) is the true total infected population. If testing of newly infecteds occurs after a distribution *ρ*(*τ* ; *n, γ*) of infection times, 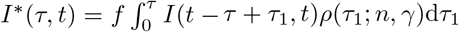

In our development of 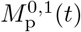 and CFR_d_(*t, τ*_res_) in the manuscript, we assumed the entire infected population was tested and confirmed. Thus, 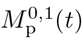 and CFR_d_(*t, τ*_res_) were computed using *f* = 1 and more accurately represent the mortality ratios of the population *conditioned* on being tested positive.

**FIG. S4.**
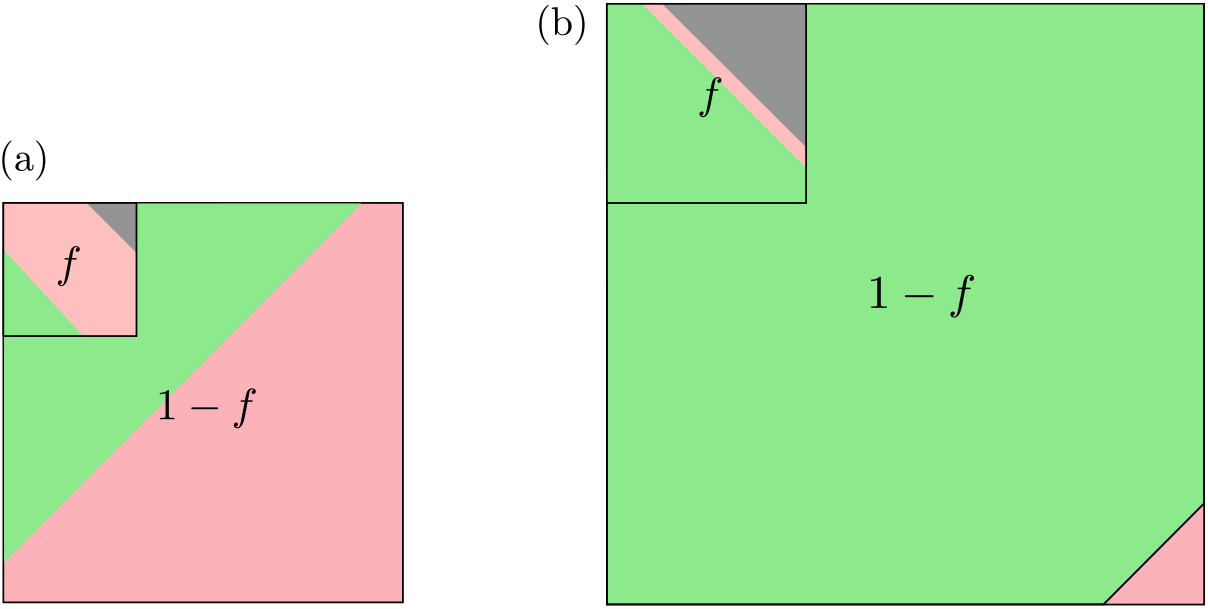
Fractional testing. An example of fractional testing in which a fixed fraction *f* of the real total infected population is assumed to be tested. The remaining 1 − *f* proportion of infecteds are untested. Equivalently, if the total tested fraction has unit population, then the total population of the untested pool is 1*/f*− 1. (a) At short times after an outbreak, the known tested infected population has not yet resolved and is composed of deaths (gray), recovereds (green), and infecteds (red). We assume that the untested fraction of infecteds (red) have mild or no symptoms, do not die, and can only recover (green). (b) At longer times, the infecteds further resolve. The *M*_p_(*t*) and CFR metrics that are based on only the tested fraction will overestimate the true mortality fraction of all infected cases.

To estimate the mortality ratio of the population conditioned simply on being infected, we have to estimate the larger number of recovereds that went untested. As shown in Fig. S4, the untested recovered fraction can be estimated by assuming that the death rate for the untested infecteds is zero and by writing an SIR model without death for the untested pool of infected

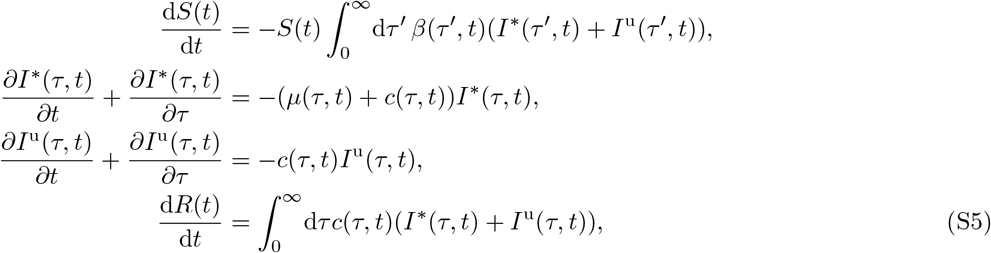

where *I*(*τ, t*) = *I*^*∗*^(*τ, t*) + *I*^u^(*τ, t*). The true mortality ratio is then straightforwardly defined by, for example,

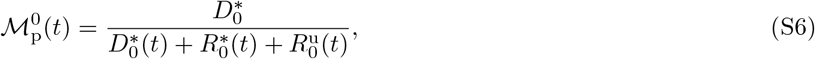

Where

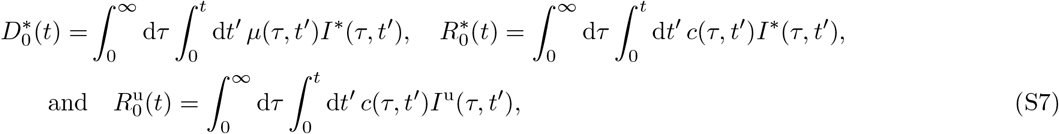

with analogous expressions for 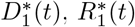, and 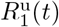. At long times, after resolution of all infecteds, the untested recovered population is

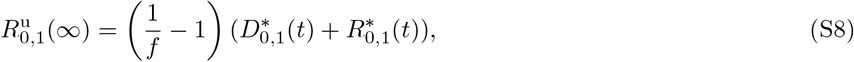

which yields the asymptotic true ratio 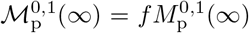 as described in the Discussion and Summary. In this simple rescaling to account for untested populations, we have assumed that all deaths come from the tested pool and that the recovery rate *c* is the same in the tested and untested pools.

### D. Influence of different transmission rates

In Fig. 3 of the main text, we observe that the population-level mortality ratio 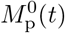 approaches a plateau during the initial exponential growth phase of an epidemic (*i*.*e*., for *S*(*t*) ≈ *S*_0_). If the number of new infections decreases (*e*.*g*., due to quarantine measures), 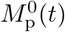 starts growing until it reaches its asymptotic value 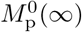 Interestingly, the pre-asymptotic values of 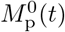 are smaller for larger infection rates *β*_1_ (see Fig. S5(a)). This counter-intuitive effect arises because larger values of *β*_1_ generate relatively larger numbers of new infected which have a lower chance of dying before *τ*_inc_ (see Eq. (4) in the main text). A similar effect occurs for non-delayed transmission (*i*.*e*., *τ*_*β*_ ≈ 0).

**FIG. S5.**
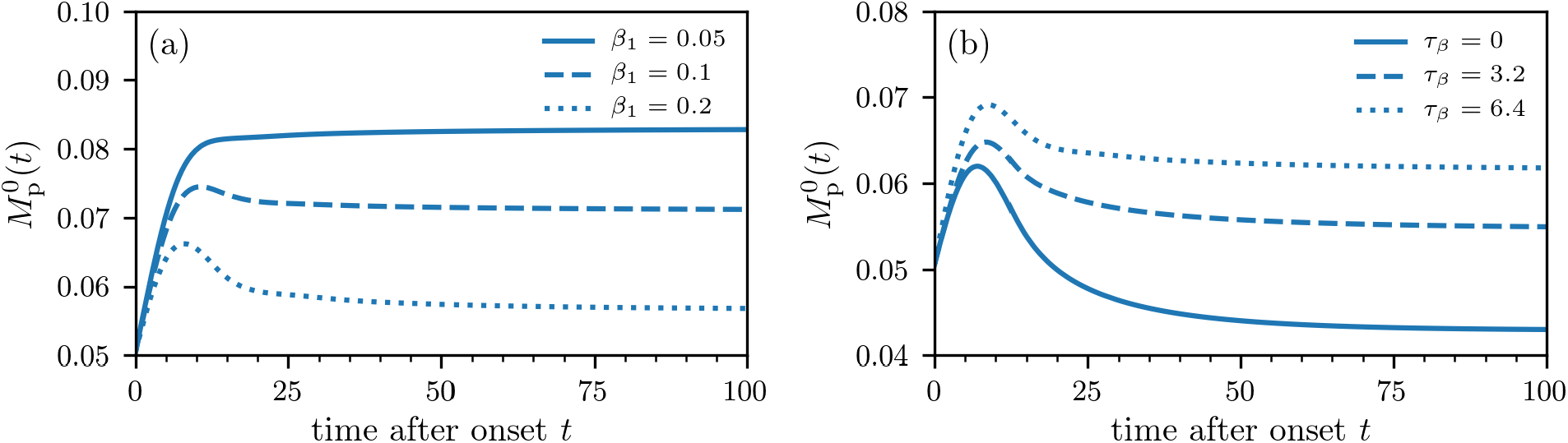
Population-level mortality for different infection rates. (a) The population-level mortality ratio 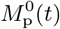 for different values of *β*_1_ and an incubation time of *τ*_inc_ = 6.4 days. In the initial exponential growth phase of the epidemic (*i*.*e*., *S*(*t*) ≈ *S*_0_), larger infection rates *β*_1_ lead to smaller values of 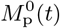. (b) We observe a similar effect for non-delayed transmissions (*i*.*e*., *τ*_*β*_ ≈ 0). As long as *S*(*t*) ≈ *S*_0_, smaller transmission delays *τ*_*β*_ lead to larger relative numbers of new infections and smaller 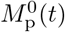.

As the transmission delay decreases, more secondary cases will result from one infection, leading to smaller values of 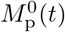 in the initial exponential growth phase of an epidemic (see Fig. S5(b)).

